# Massively parallel identification of functionally consequential noncoding genetic variants in undiagnosed rare disease patients

**DOI:** 10.1101/2021.11.02.21265771

**Authors:** Jasmine A. McQuerry, Merry Mclaird, Samantha N. Hartin, John C. Means, Jeffrey Johnston, Tomi Pastinen, Scott T. Younger

## Abstract

Clinical whole genome sequencing has enabled the discovery of potentially pathogenic noncoding variants in the genomes of rare disease patients with a prior history of negative genetic testing. However, interpreting the functional consequences of noncoding variants and distinguishing those that contribute to disease etiology remains a challenge. Here we address this challenge by experimentally profiling the functional consequences of rare noncoding variants detected in a cohort of undiagnosed rare disease patients at scale using a massively parallel reporter assay. We demonstrate that this approach successfully identifies rare noncoding variants that alter the regulatory capacity of genomic sequences. In addition, we describe an integrative analysis that utilizes genomic features alongside patient clinical data to further prioritize candidate variants with an increased likelihood of pathogenicity. This work represents an important step towards establishing a framework for the functional interpretation of clinically detected noncoding variants.

## Introduction

The application of whole genome sequencing (WGS) in a clinical setting has greatly facilitated the discovery of disease-associated genetic variants, particularly rare noncoding variants that occur outside of protein-coding genes. Noncoding variants can be pathogenic in cases where they disrupt the activity of important functional regulatory elements^1,2^. However, in most cases the functional consequences of a rare noncoding variant cannot be predicted based on sequence alone. As a result, the expanded variant identification achieved with WGS has had a limited impact on diagnostic rates in the clinic, which have remained below 50%^3–5^.

At present, there are limited clinical practices for distinguishing pathogenic noncoding variants. Numerous *in silico* approaches have been developed for predicting noncoding variant pathogenicity based on a variety of genomic features. Nearly all these methods utilize evolutionary parameters to infer pathogenicity^6,7^. A subset of these tools also incorporates molecular features such as chromatin accessibility and transcription factor binding profiles^8,9^. Alternative prediction methods rely more heavily on population frequencies and/or previously reported disease associations^10,11^. Unfortunately, these computational approaches have low concordance in their predictions of pathogenicity and often contradict experimental data^12^.

Experimental tools have been developed for profiling the biological activity of noncoding genomic sequences at scale in cell-based models. More specifically, the massively parallel reporter assay (MPRA) is a high-throughput sequencing-based approach that combines array-based DNA synthesis with a plasmid-based reporter system and permits the simultaneous quantitative assessment of regulatory capacity for thousands of noncoding sequences of interest^13,14^. The MPRA has previously been used to explore the functional impact of noncoding variants reported by the 1000 Genomes Project and disease-associated noncoding variants identified in genome-wide association studies (GWAS)^15,16^. Although the MPRA has proven to be a powerful tool for characterizing noncoding genomic sequences, it has yet to be applied towards variant interpretation in a clinical setting.

Here, we describe the implementation of an MPRA-based strategy for characterizing the functional consequences of rare noncoding variants detected directly from whole genome sequencing of rare disease patients with a history of negative genetic testing. We profiled >3,000 genomic regions of interest and identified hundreds that exhibit significant regulatory activity. Furthermore, we illuminated >100 rare noncoding variants that significantly alter the regulatory capacity of these genomic regions. Importantly, the functional consequences of these variants could not have been predicted in the absence of experimental data.

In addition to profiling the impact of rare noncoding variants on regulatory activity we outline an integrative approach for prioritizing candidate variants with an increased likelihood of pathogenicity. We incorporate transcription factor binding profiles at variant sites alongside clinical data from each proband to strengthen phenotypic associations between variant-containing regulatory elements and clinical presentations. Using this method we uncover several consequential variants that occur within genomic contexts that may contribute to disease etiology. In summary, this study provides a roadmap for the implementation of MPRAs in a clinical setting and introduces an important tool for improving the interpretation of noncoding variants in rare disease genomes.

## Results

### Identification of rare noncoding genetic variants in a cohort of undiagnosed rare disease patients

To explore the impact of rare noncoding genetic variants on the regulatory potential of genomic sequences we first identified rare variants in a cohort of undiagnosed rare disease patients. We analyzed the genomes of 111 exome negative probands, an even mix of females and males with a median age of 7 years at the time of analysis (**Figures 1a, 1b**). The patient cohort presented with a wide variety of clinical manifestations spanning all organ systems (**Figure 1c**). Among the most prevalent clinical features were anomalies of the nervous system, the musculoskeletal system, and the cardiovascular system (**Figure 1c**).

**Figure 1.**
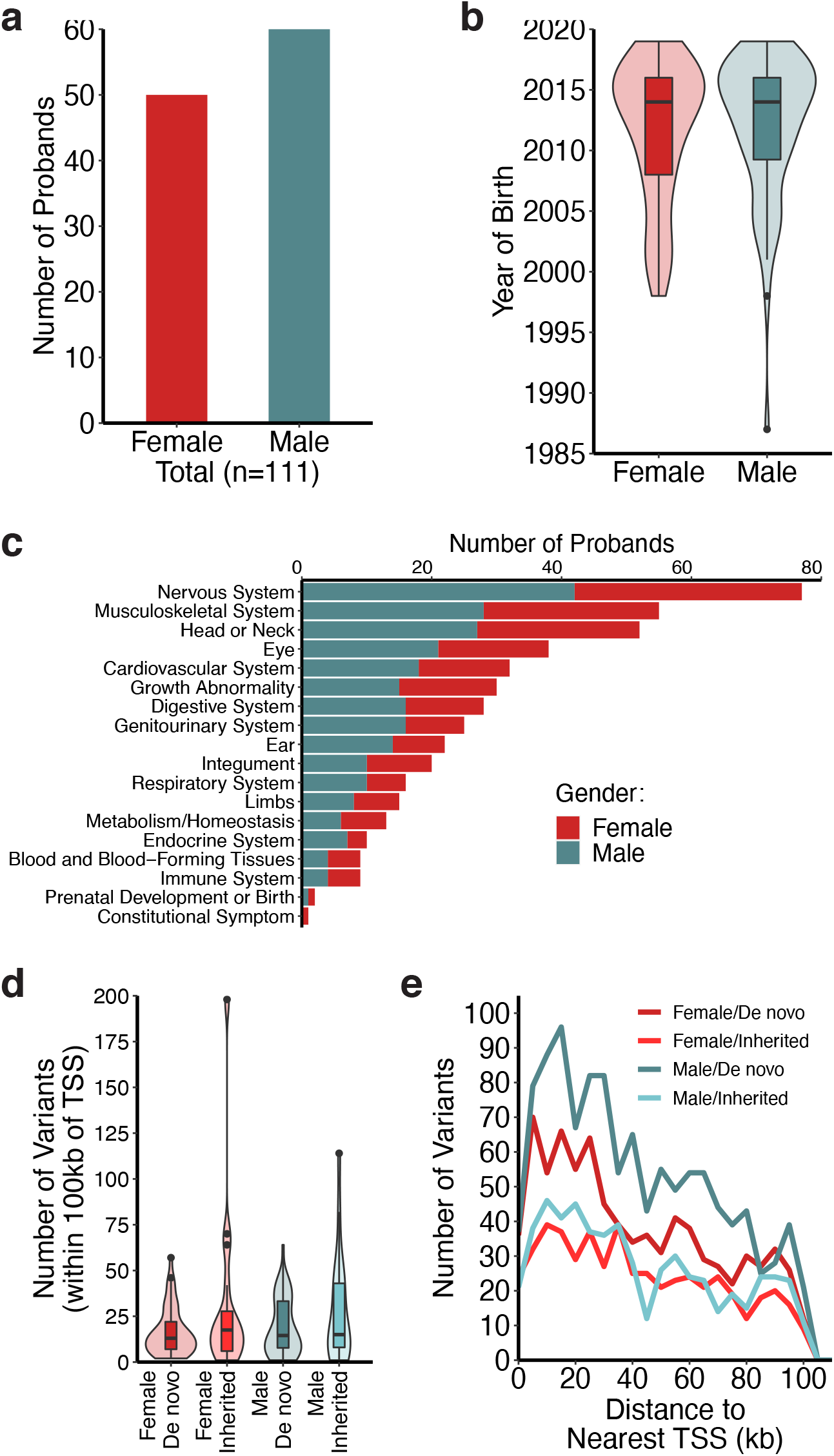
Identification of rare genetic variants from an undiagnosed rare disease cohort. **a** Gender composition of proband cohort.**b** Age distribution of proband cohort. **c** Prevalence of clinical features associated with selected patients. **d** Frequency of rare variants detected within 100 kb of a TSS. **e** Distance to nearest TSS for detected rare variants.

For this study we focused exclusively on rare noncoding single nucleotide variants occurring within 100kb of an annotated transcription start site. Variants were designated as rare if they had a gnomAD minor allele frequency < 0.001 (internal allelic frequencies were used when gnomAD data was not available) and were detected using at least two independent sequencing technologies. Family trio sequencing was available for all probands in this study, allowing us to further distinguish *de novo* rare variants from those inherited from a parent. We observed a median of 14 *de novo* rare variants per proband within 100kb of a TSS and a total of 1,958 *de novo* variants within this genomic window across all 111 probands (**Figure 1d, Table S1**). For comparison we also selected inherited rare variants from a subset of probands. Inherited variants were selected at a similar frequency, a median of 15.5 variants per proband within 100kb of a TSS, for a total of 1,101 inherited rare variants (**Figure 1d, Table S1**). In the probands profiled in this study we found that *de novo* rare variants were detected throughout the +/-100kb genomic window surrounding TSSs and were slightly more prevalent in genomic regions near TSSs (**Figure 1e**).

### Profiling the regulatory capacity of genomic sequences using a massively parallel reporter assay

To systematically evaluate the regulatory potential of rare variant-containing genomic sequences we designed a massively parallel reporter assay (MPRA). For our MPRA we designed a library of sequences composed of 100 nucleotides of genomic information centered around the chromosomal position of each rare noncoding variant we detected (**Figure 2a, Table S1**). We designed four analogous sequences for each genomic region of interest to represent all possible nucleotides in the variant position (**Figure 2a, Table S1**). These sequences were used to design an oligonucleotide library with each oligo containing a genomic sequence of interest as well as a unique 12 nucleotide barcode sequence. We assigned each genomic sequence of interest 5 independent barcodes resulting in an overall library size of 61,180 oligos (3,059 variants x 4 sequence analogs per variant x 5 barcodes per sequence) (**Table S1**).

**Figure 2.**
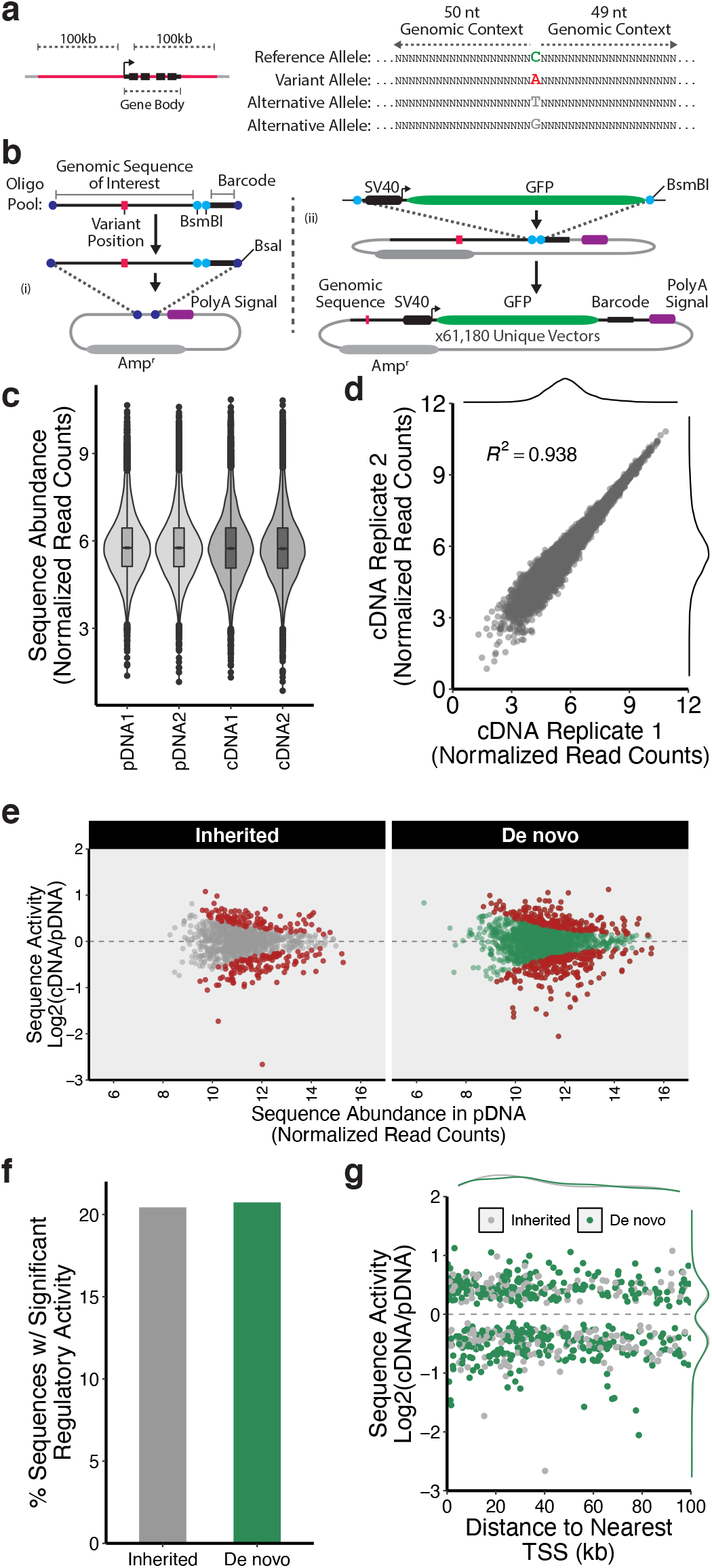
Profiling the regulatory capacity of genomic sequences using a massively parallel reporter assay. **a** Schematic of MPRA oligonucleotide library design. **b** Schematic of MPRA vector design and cloning. **c** Abundance of MPRA library elements corresponding to genomic sequences represented in the MPRA plasmid pool (pDNA, technical replicates) and expressed in HEK293T (cDNA, biological replicates). **d** Reproducibility of MPRA library element detection across biological replicates. **e** Regulatory activity of reference genome sequences profiled in MPRA (red = significant regulatory activity). **f** Fraction of reference genome sequences that display significant regulatory activity in MPRA. **g** Distance to nearest TSS for reference genome sequences that display significant regulatory activity.

We next constructed an MPRA plasmid library using a two-step cloning approach. First, the oligo library was cloned into an empty plasmid (no reporter gene) backbone (**Figure 2b**). In the second cloning step a reporter cassette containing an SV40 promoter followed by GFP was inserted between the genomic sequence of interest and its associated barcode sequence (**Figure 2b**). In the final MPRA library pool each plasmid encodes a uniquely barcoded GFP transcript that permits the association of its expression with the upstream genomic sequence of interest (**Figure 2b**). Following library construction we performed targeted sequencing of the barcode containing region of the plasmid pool and confirmed that all of the genomic sequences of interest were represented in the MPRA plasmid library (**Figure 2c, Table S2**).

To measure the regulatory potential of the genomic sequences in our MPRA library we transfected the plasmid pool into HEK293T cells. Given the wide array of clinical manifestations in the patient cohort, we reasoned that potentially pathogenic noncoding variants would exhibit measurable effects on reporter expression in most cellular contexts. We chose HEK293T as a cellular system due to the cell type’s robust growth properties and high transfection efficiency. Cells were harvested 24 hours after transfection and targeted RNA-Seq libraries were generated by amplifying the barcode-containing region of the expressed GFP reporter transcripts. We detected robust expression of reporter transcripts associated with each of the genomic sequences of interest in our MPRA library (**Figure 2c, Table S2**). Moreover, we observed high concordance in reporter expression associated with each respective sequence of interest across biological replicates (**Figure 2d**).

In the MPRA experiment the abundance of each barcode in the RNA-Seq library serves as a proxy for the regulatory potential of the associated genomic sequence of interest. To establish a baseline for regulatory activity we first profiled MPRA expression for library elements representing the reference genome. After normalizing library element abundance in the RNA-seq expression library to abundance in the MPRA plasmid pool we found that many reference genome sequences were capable of significantly influencing reporter expression (**Figure 2e, Table S2**). Overall, we observed significant regulatory activity from ∼20% of the reference genome sequences in our MPRA library (**Figure 2f**). Sequences displaying significant regulatory activity were distributed evenly between genomic regions corresponding to inherited or *de novo* variants identified in our patient cohort (**Figures 2e, 2f**). Moreover, we found that the regulatory activity of these sequences was split evenly between activation and repression of reporter expression (**Figure 2e**). Interestingly, regulatory activity in the MPRA showed no correlation with the distance between the genomic sequence of interest and its nearest TSS in the genome (**Figure 2g**). Altogether, these results demonstrate that the MPRA is a robust method for profiling the regulatory activity of genomic sequences.

We next searched for MPRA library element features that might be associated with regulatory activity. In general, we observed uniform regulatory activity from genomic sequences representing variant-containing regions across all probands in our patient cohort (**Figure S1a**). Furthermore, we detected uniform activity across the sequence analogs we designed for each variant (**Figure S1b**). Expression in the MPRA was not influenced by the identity of the nucleotide in the variant position (**Figure S1c**). Likewise, expression was similar across sequences corresponding to genomic regions containing inherited or *de novo* variants (**Figure S1d**). Lastly, genomic features such as chromatin accessibility at the variant site were not predictive of regulatory activity (**Figure S1e**). These data highlight the ability of the MPRA to discern sequences with regulatory capacity in the absence of predictive features.

### Quantifying the impact of genetic variants on the regulatory capacity of genomic sequences

To evaluate the impact of rare genetic variants on the regulatory capacity of the genomic sequences we compared expression between corresponding reference and variant alleles represented in our MPRA library. We found that ∼4.5% of the variants profiled had a significant impact on regulatory activity (**Figures 3a, 3b, Table S3**). Consequential variants were distributed evenly between genomic regions harboring inherited or *de novo* variants (**Figure 3b**). Furthermore, there was no relationship between the class of base mutation (transition or transversion) and the effect of the variant on regulatory activity (**Figure S2a**). Similar to the basal regulatory activity detected from reference alleles, we observed no correlation between the functional impact of a variant and its distance from the nearest TSS in the genome (**Figure 3c**).

**Figure 3.**
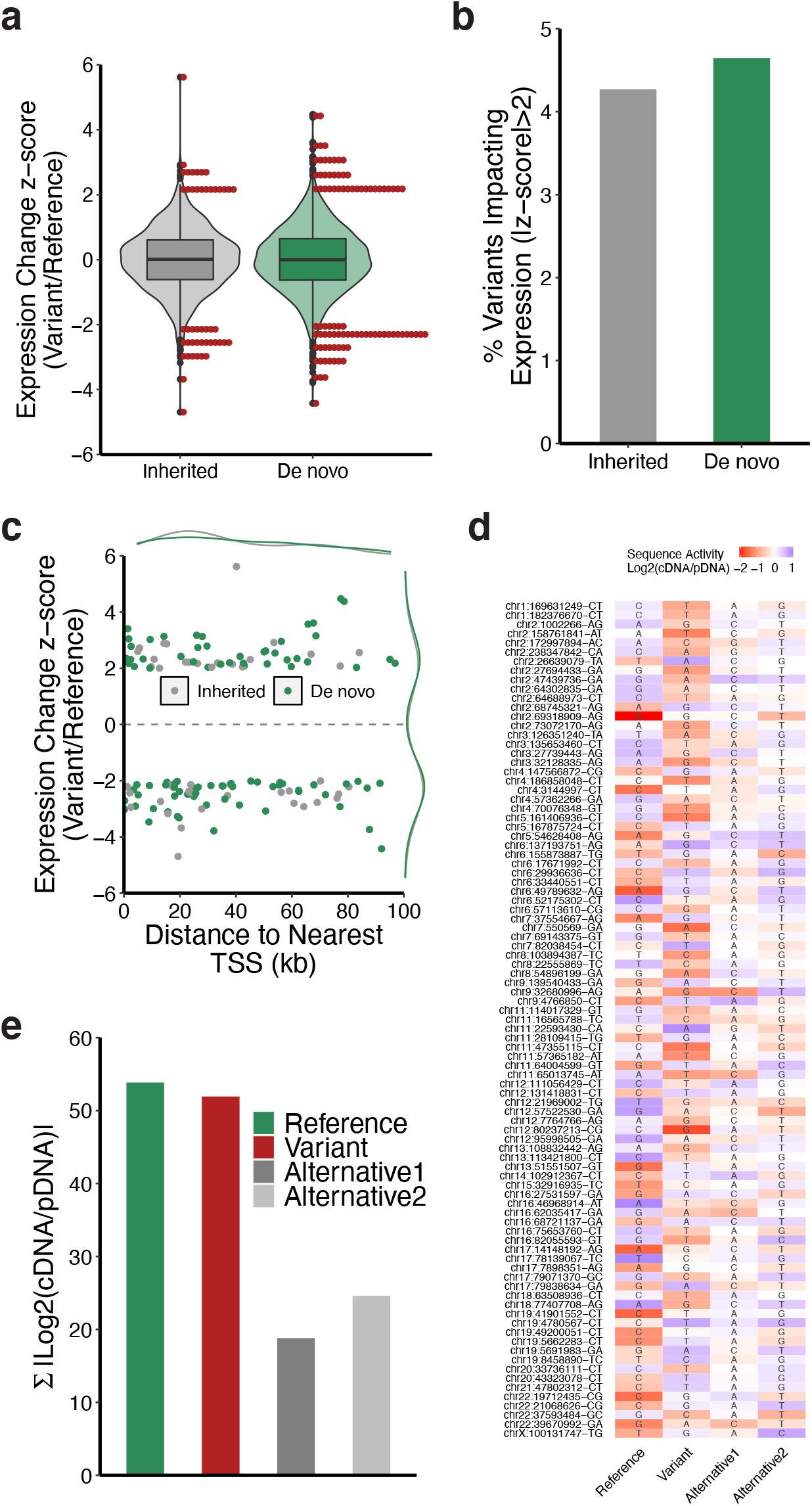
Rare genetic variants alter the regulatory capacity of genomic sequences. **a** Distribution of expression differences between reference and variant alleles profiled in MPRA (red points denote variants with an expression difference |z-score| > 2). **b** Fraction of profiled rare variants that alter regulatory capacity of genomic sequences. **c** Distance to nearest TSS for variants that alter the regulatory capacity of genomic sequences (|z-score| > 2). **d** Heatmap showing regulatory capacity of all possible alleles corresponding to *de novo* variants that display altered regulatory capacity (|z-score| > 2). **e** Aggregated regulatory capacity of genomic sequences shown in **3d** by allele type.

Overall, we identified 91 *de novo* variants that significantly impacted the regulatory potential of genomic sequences (**Figure 3d, Table S3**). We detected 51 *de novo* variants that resulted in significant decreases in reporter expression and 40 *de novo* variants resulting in significant increases in reporter expression. Importantly, increases in reporter expression appeared to result from a loss of repression observed by the reference allele (**Figure 3d**). Analysis of MPRA expression from all possible alleles corresponding to *de novo* variants that significantly alter reporter expression revealed that most of the regulatory activity was associated with reference or variant alleles and that other alternative alleles had minimal regulatory potential (**Figures 3d, 3e**). These results demonstrate that the MPRA is a powerful tool for identifying genetic variants that impact the regulatory capacity of genomic sequences.

### Integrative prediction of rare variant pathogenicity

To illuminate rare variants with a higher likelihood of pathogenicity we integrated human phenotype ontology (HPO) terms into our MPRA analysis. Briefly, HPO provides a standardized terminology for phenotypic features associated with human disease^17^. As a resource, HPO also documents associations between HPO terms and genes with demonstrated links to annotated phenotypes. We reasoned that a rare variant occurring within a regulatory element could result in phenotypes similar to those associated with transcription factors bound to the element. To test this hypothesis we utilized publicly available ChIP-seq data to assess transcription factor binding in the genomic regions profiled by our MPRA library. We subsequently evaluated the overlap in HPO terms associated with each transcription factor and the terms associated with probands harboring variants at binding sites for each factor. We identified 145 variants (61 inherited and 84 *de novo*) for which there was significant overlap in HPO terms between the proband and at least one transcription factor bound to the variant-containing site (**Figure 4a, Table S4**). Interestingly, variants that had significant overlap in HPO terms with bound transcription factors as well as a significant impact on regulatory activity in our MPRA were almost exclusively *de novo* (**Figure 4a**).

**Figure 4.**
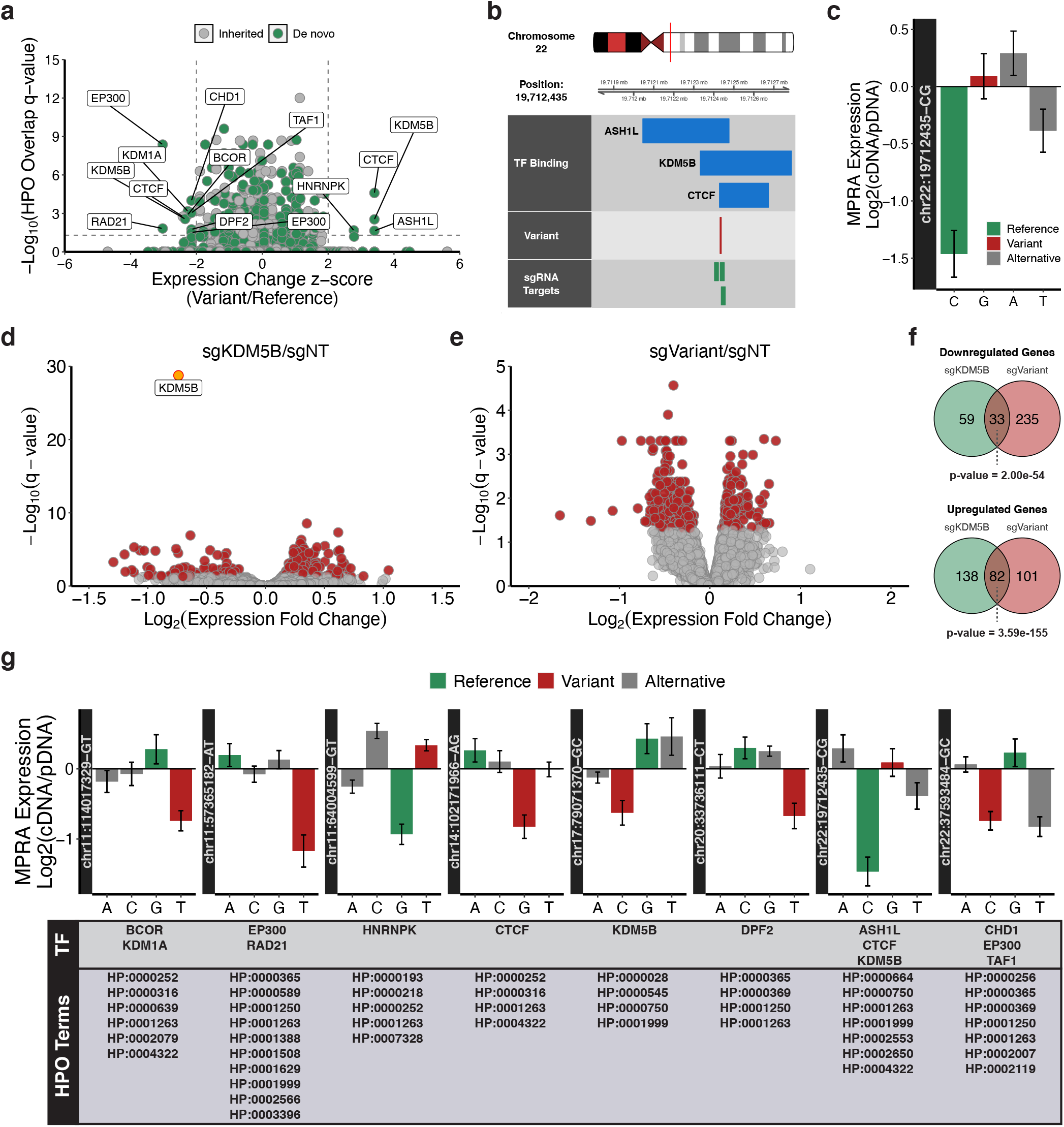
Integrative prediction of rare variant pathogenicity. **a** Significance analysis of HPO term overlap between probands and variant-associated transcription factors. **b** Schematic of transcription factor binding at candidate variant position. **c** MPRA analysis of candidate variant. **d** Volcano plot showing transcriptome-wide expression changes following CRISPRi-mediated repression of KDM5B. **e** Volcano plot showing transcriptome-wide expression changes following CRISPRi-mediated repression of variant site. **f** Significance analysis of overlapping expression changes following KDM5B or variant site repression. **g** MPRA analysis, transcription factor binding, and overlapping HPO terms for candidate variants.

We next evaluated whether transcription factor binding profiles can be predictive of the phenotypic consequences of noncoding variants. We selected a candidate *de novo* variant on chromosome 22 detected in a proband that had significant HPO term overlap with several factors that bind to the variant site (**Figure 4b**). Expression analysis in the MPRA demonstrated that the reference allele corresponding to this variant exhibited repressive features that were absent with the variant allele (**Figure 4c**). In agreement with this observation, one of the factors that binds to this site (KDM5B) is a lysine demethylase known to play a role in transcriptional repression^18^. Based on this relationship we predicted that inhibiting the function of KDM5B and perturbing the putative regulatory activity of the genomic region harboring the variant would have overlapping consequences.

To explore a potential association between KDM5B and the variant-containing genomic region we inhibited expression of KDM5B using CRISPR interference (CRISPRi) and profiled the downstream effects using transcriptome-wide expression analysis. In response to KDM5B inhibition we detected significant expression changes in 312 genes (220 upregulated, 92 downregulated) (**Figure 4d, Table S5**). In parallel to KDM5B inhibition we used CRISPRi to interfere with the potential regulatory functions of the variant-containing genomic region which resulted in significant expression changes in 451 genes (268 upregulated, 183 downregulated) (**Figures 4b, 4e, Table S5**). Importantly, the large number of differentially expressed genes in response to variant site perturbation indicates that the genomic region harboring the variant is a functional regulatory element. We observed significant overlap in the genes that were differentially expressed in response to KDM5B inhibition and variant site perturbation suggesting the presence of regulatory interactions between the transcription factor and the variant site (**Figure 4f**). These results are consistent with the overlap in phenotypes (HPO terms) associated with KDM5B and the proband harboring the variant and support the use of transcription factor binding profiles for predicting the consequences of noncoding variants.

Overall, we identified 8 rare variants (7 *de novo*, 1 inherited) that exhibited a significant impact on reporter expression in our MPRA and occurred in a genomic region bound by transcription factor(s) associated with HPO terms that overlapped the respective proband (**Figure 4g**). Our MPRA expression data revealed that 6 of these variants were loss-of-function (decreased expression) and 2 variants were gain-of-function (loss of repression). Although the 7 *de novo* variants that ultimately met our stringent prioritization criteria represent just 0.36% (7/1958) of the total *de novo* variants in the MPRA library, those variants represent 8.33% of the *de novo* variants that occurred in genomic regions bound by transcription factor(s) with overlapping HPO terms. This observation suggests that incorporating transcription factor binding profiles into the design of future MPRA libraries will likely lead to more efficient identification of high priority variants. Altogether, our results demonstrate that the MPRA, in combination with patient clinical data and complementary genomic data, is an effective method for illuminating rare noncoding variants that warrant detailed investigation.

## Discussion

A growing body of evidence has demonstrated that most disease-associated variants occur in noncoding regions of the genome^19–21^. Although several computational tools have been developed for predicting the pathogenicity of noncoding variants, classifications from these tools exhibit low concordance and no single approach has been adopted as standard practice in the clinical interpretation of noncoding variants^12^. For rare noncoding variants that have not been previously detected or characterized, experimental evidence is nearly essential to evaluate the functional impact of the variant prior to consideration for involvement in disease. Here, we provide a scalable solution to this challenge by applying high-throughput functional genomic technologies to assist with the interpretation of clinically detected noncoding variants. We constructed an MPRA to directly profile the functional consequences of thousands of rare noncoding variants detected in the genomes of undiagnosed rare disease patients. Furthermore, we have outlined an integrative prioritization strategy that incorporates patient clinical data along with publicly available genomic data to pinpoint noncoding variants with an increased likelihood of pathogenicity.

Our MPRA-based strategy enabled us to systematically profile the functional consequences of 3,059 clinically detected rare noncoding variants and our analysis pipeline led to the prioritization of 8 noncoding variants for follow up study. Although the overall yield of prioritized variants in this study was relatively low, our data have revealed features of the MPRA design that can be adapted to significantly improve the effectiveness of future screening campaigns. For example, we found that nearly all the noncoding variants prioritized using our approach were *de novo*. Furthermore, profiling all possible alleles at each variant site did not generate insight that would have a meaningful impact on variant interpretation. By focusing future MPRA library designs exclusively on *de novo* variants, profiling only reference or variant alleles, and further restricting libraries to genomic regions bound by transcription factors associated with relevant HPO terms we estimate that a 30-fold increase in the volume of prioritized variants could be achieved. Such an improvement would significantly increase the number of pathogenic variants uncovered using this approach.

The variants we profiled in this study were identified in rare disease patients that exhibited a variety of clinical manifestations spanning multiple organ systems suggesting that the effects of potentially pathogenic variants are not restricted to specific cellular contexts. However, this is unlikely to be the case for all patients that undergo WGS and defined cellular models may be required to profile variants associated with isolated clinical presentations. Aside from cellular context, there are additional aspects of MPRA design that are likely to impact experimental results. While the MPRA we describe in this study was performed using episomal expression vectors, lentiviral-based MPRA systems that integrate into the genome have been developed^22,23^. Chromosomally integrated MPRAs may be required to profile variants with functional consequences that are dependent on chromatin context.

Among the most critical elements of the approach we outline in this study is direct access to patient clinical data. This information is essential for establishing phenotypic associations between genomic regions that harbor noncoding variants and clinical features of disease. The utility of patient data when predicting the phenotypic consequences of noncoding variants further supports the implementation of our strategy directly within a clinical setting. Although the approach we describe here is intended to facilitate the clinical interpretation of noncoding variants at the level of individual patients, data sharing across the broader research community is of fundamental importance to the process of rare disease diagnosis. Importantly, any functional data obtained when profiling a rare disease genome has the potential to aid in the interpretation of unrelated disease genomes that harbor variants in the same chromosomal positions or in proximal genomic regions.

In summary, this study provides proof of principle for the application of an MPRA-based strategy to assist with the clinical interpretation of noncoding variants identified in disease genomes. We anticipate that such an approach will have a positive impact on the diagnostic rates achieved by clinical WGS.

## Methods

### Selection of candidate rare variants

Single nucleotide variants were called using DRAGEN 3.6.3 on GRCh38 for probands and parents enrolled in the Genomic Answers for Kids initiative at Children’s Mercy Hospital. The following criteria were used to select candidate variants from probands: a) gnomAD minor allele frequency <0.001 (internal allelic frequencies were used when gnomAD data was not available), b) variant detection using at least two sequencing technologies (whole genome sequencing, whole genome bisulfite sequencing, 10X linked-read sequencing, or PacBio long-read sequencing), and c) variants occurring within 100kb of an annotated transcription start site. Variants were excluded from consideration if: a) observed in more than one proband, b) previously reported as causative of a disorder, or c) predicted to impact protein-coding gene function through nonsense mutation, frameshift mutation, disruption of a stop codon, loss of translation initiation, or interference with splice donor/acceptor sequences. Candidate rare variants were categorized as “inherited” if observed in sequencing reads from parental sequencing. Candidate rare variants were categorized as “*de novo*” if no evidence of the variant was present in parental sequencing data. Variant coordinates were lifted over to GRCh37 for genomic analyses.

### MPRA library cloning

The MPRA oligo library was amplified using Q5 High-Fidelity DNA Polymerase (NEB). The amplified library was size-selected using Agencourt AMPureXP beads (Beckman Coulter). The oligo library was then inserted into an empty MPRA vector by golden gate assembly using BsaI (NEB) and T4 Ligase (NEB). The resulting library was purified using isopropanol precipitation (15 µl elute) then expanded by electroporation into 5 vials (3 µl ligation/vial) of One Shot TOP10 Electrocomp E. coli (ThermoFisher). The plasmid library was isolated using the Plasmid Plus Maxi kit (QIAGEN). The MPRA reporter gene (SV40 promoter followed by GFP) was then incorporated into the plasmid library by golden gate assembly using Esp3I (NEB) and T4 Ligase (NEB). The final MPRA plasmid library was purified, expanded, and isolated as described previously. Primers and PCR conditions are listed in Supplemental Table S6.

### Cell Culture

HEK293T cells (ATCC) were cultured in Dulbecco’s Modified Eagle’s Medium (DMEM) supplemented with 10% fetal bovine serum (Life Technologies) and 1% Penicillin-Streptomycin (ThermoFisher).

### Lentivirus Production

HEK293T cells (ATCC) were seeded in a 6-well dish at a density of 1×10^6^ cells per well. Twenty-four hours after plating cells were transfected with a mixture of 8.25 µL TransIT LT-1 reagent (Mirus Bio) plus 66.75 µL Opti-MEM (Gibco) to which 1250 ng psPAX2 vector DNA (Addgene 11260), 125 ng pMD2.G vector DNA (Addgene 11259) and 1250 ng pLX_311-KRAB-dCas9 vector DNA (Addgene 96918) were added. After a 30 minute incubation at room temperature, this mixture was applied to cells. Lentiviral supernatant was collected 48 h post-transfection.

### Generation of stably-expressing dCas9-KRAB HEK293T cell line

To generate a stably-expressing dCas9-KRAB line, 1 mL lentivirus was applied to 5×10^5^ HEK293T cells with the addition of 6 µg/mL polybrene (Sigma). Virus-containing medium was replaced with fresh DMEM supplemented with 10% FBS at 24 h post-infection. At 48 h post-infection, 8 µg/mL blasticidin (Gibco) was applied to begin selection. Cells were used for downstream CRISPR interference experiments following at least 72 h of selection, at which time all uninfected control cells were dead.

### Single guide RNA cloning

Forward and reverse-complement single-stranded oligonucleotide inserts containing 5’ BsmBI sites followed by single guide RNA sequences were purchased from IDT. Oligos were mixed at equimolar concentrations in NEB Buffer 3.1 and annealed using thermal cycler conditions listed in Supplemental Table S6. Following annealing of complementary oligonucleotide inserts, inserts were ligated into BsmBI-digested pXPR_050 vector (Addgene cat. no. 96925) using the Quick Ligation Kit (NEB) according to manufacturer’s protocol and the ligation product was transformed into Stbl3 chemically competent E. coli cells (Invitrogen) via heat shock. Sequence-confirmed clones were cultured and pDNA extracted and purified using the Plasmid Plus Midi Kit (Qiagen).

### CRISPR interference

HEK293T cells stably-expressing dCas9-KRAB were plated in 6-well dishes at a density of 2×10^5^ cells/well. Cells were transfected 24 h after plating with an equimolar pool of 3 plasmids encoding sgRNAs targeting a common gene or variant. Cells were transfected using the Lipofectamine 3000 kit (Invitrogen) according to manufacturer directions. Briefly, a mixture of 5 µL P3000 reagent, 833 ng of each of the 3 sgRNA-containing pXPR_050 vectors targeting the same gene or variant, and 125 µL Opti-MEM was added to a mixture of 7.5 µL of Lipofectamine 3000 and 125 µL of Opti-MEM. The mixture was allowed to incubate at room temperature for 15 minutes, after which it was added to cells in a drop-wise manner. RNA was isolated 72 h post-transfection using the RNEasy mini kit (Qiagen) as per the manufacturer’s instructions.

### RNA Sequencing and differential expression analysis

RNA sequencing libraries were prepared with the TruSeq Stranded Total RNA Library Prep Gold Kit (Illumina) as per the manufacturer’s instructions using 1000 ng of input RNA for each library. RNA-Seq libraries were sequenced (2×150bp paired-end) on a NovaSeq (Illumina). Sequencing reads were aligned to the human genome (hg38) using STAR with default parameters^24^. Transcript abundances were determined using featureCounts with default parameters and Gencode 38 as the reference transcriptome^25^. Differential Expression was calculated using DEseq2 with default parameters^26^.

### MPRA plasmid pool transfection

Lipofectamine 3000 (Life Technologies) was used to deliver the MPRA plasmid pool into HEK293T cells as per the manufacturer’s instructions. Cells were plated in six-well dishes (3 plates per replicate) at a density of 1.5×10^5^ cells/well. Cells were transfected with MPRA plasmid pools (1 µg/well) 24 h after plating and additional culture media was added 3-4 h post-transfection. RNA from transfected HEK293T cells was isolated 24 h post-transfection using 1mL TRIzol (Life Technologies) as per the manufacturer’s instructions. RNA was then pooled from each replicate set of 6-well dishes. For each replicate, 1 µg of RNA was reverse transcribed using Superscript III Reverse Transcriptase (Life Technologies). RNA was treated with DNase I (Worthington) prior to reverse transcription.

### MPRA targeted sequencing

MPRA targeted sequencing libraries were generated directly from 50% (10 µl) of each cDNA reaction using Q5 High-Fidelity DNA Polymerase (NEB). Libraries were size-selected using Agencourt AMPureXP beads (Beckman Coulter). Prior to sequencing, the concentration of each library was assessed using a NanoDrop One Microvolume UV-Vis Spectrophotometer (Thermo Scientific). Primers and PCR conditions are listed in Supplemental Table S6. MPRA libraries were sequenced on a NovaSeq (Illumina).

### MPRA expression analysis

MPRA targeted sequencing libraries were generated such that the first 12 bases of each read corresponded to the 12-base tag used to uniquely identify individual oligonucleotides in the MPRA library. Expression driven by each sequence-of-interest in the library was defined by the sum of the reads mapping to each of the 5 distinct tags corresponding to the respective sequence-of-interest. To evaluate regulatory activity the abundance of reads mapping to each sequence-of-interest in the cDNA libraries were compared to the abundance in the MPRA plasmid library using DESeq2.

### Human Phenotype Ontology term overlap significance

Significance analysis of HPO term overlap was calculated using the hypergeometric distribution given the number of HPO terms associated with each gene or patient under comparison and the total number of annotated HPO terms. Resulting P-values were corrected for multiple hypothesis testing using the Benjamini-Hochberg method.

### Acquisition of publicly available datasets

Human phenotype terms were obtained from the Human Phenotype Ontology^17^. Human transcription factor binding sites and DNaseI hypersensitivity profiles were obtained through the ‘Txn Factr ChIP E3 (encRefTfbsClustered)’ table and the ‘DNase Clusters (wgEncodeRegDnaseClusteredV3)’ table, respectively, from the UCSC Genome Browser^27^.

## Data Availability

The Gene Expression Omnibus accession number for the MPRA and bulk RNA sequencing data described in this paper is GSE185795. The dbGaP accession number for whole genome sequencing data described in the paper is phs002206.v2.p1.

## Acknowledgements

This work was supported by generous philanthropic contributions to the Children’s Mercy Research Institute and the Genomic Answers for Kids program.

## Authors’ contributions

STY conceived the study. JAM, MM, SNH, JCM, and STY designed and performed experiments. JJ, TP, and STY performed computational analyses. STY wrote the manuscript with assistance from all authors. STY supervised the study. All authors read and approved the final manuscript.

## Figure Legends

**Figure S1.**
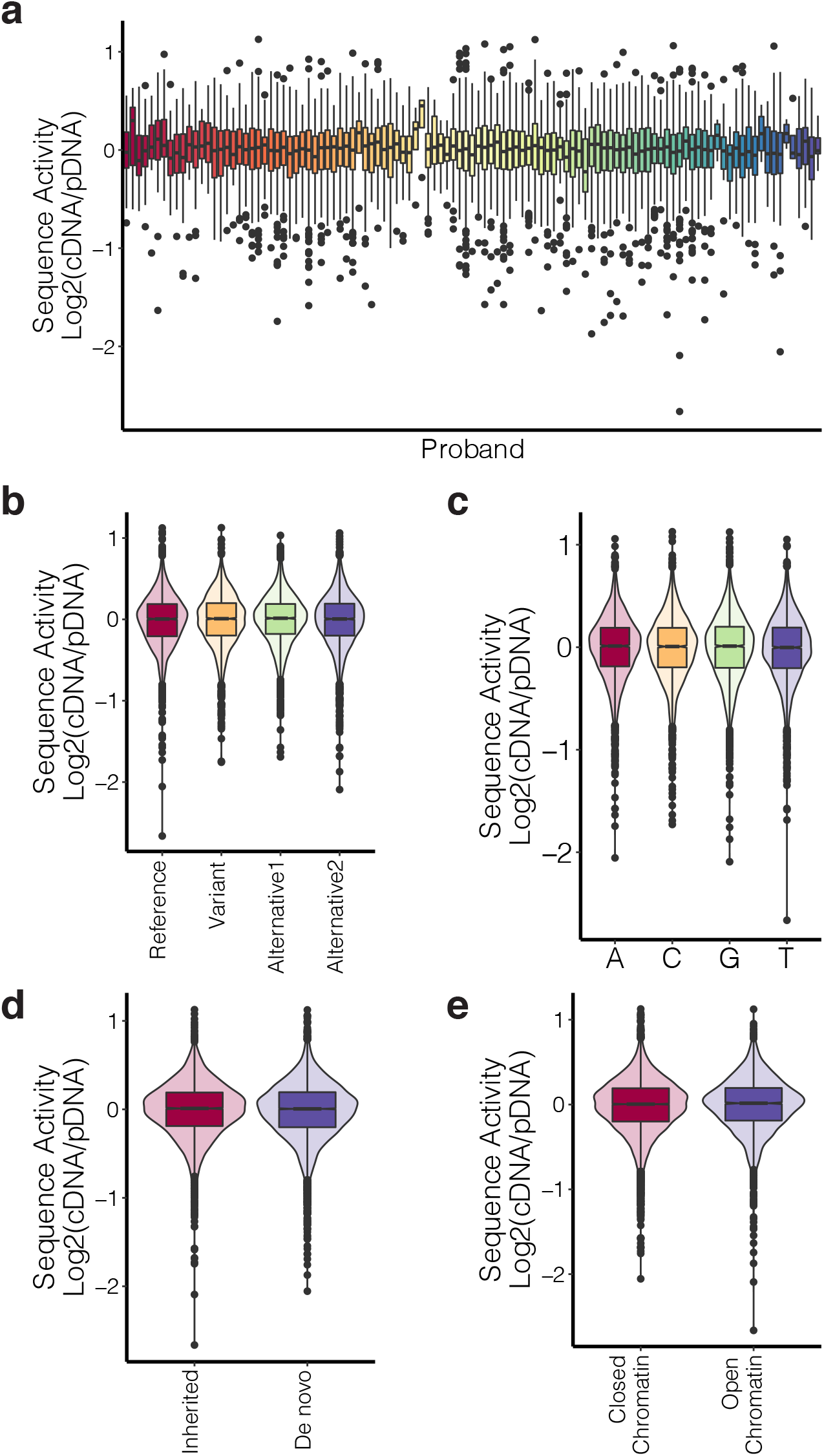
MPRA sequence activity does not correlate with basic sequence features. **a-e** Distribution of regulatory activity from genomic sequences profiled in MPRA separated by proband (**a**), allele type (**b**), base (**c**), inheritance type (**d**), and chromatin accessibility as determined by DNase I hypersensitivity (**e**).

**Figure S2.**
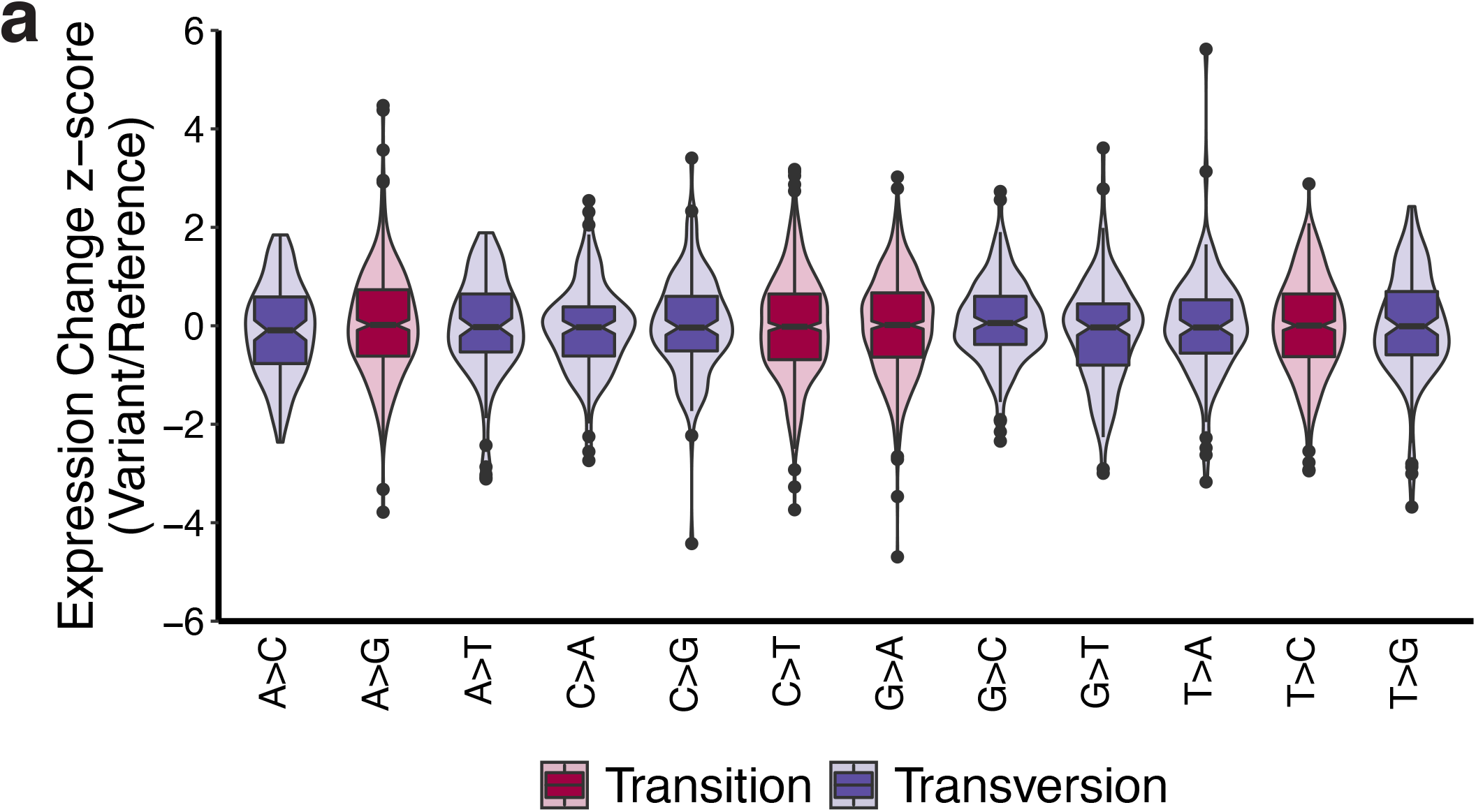
MPRA expression changes do not correlate with type of base mutation. **a** Distribution of expression differences between reference and variant alleles profiled in MPRA separated by type of base mutation.

**Table S1. MPRA library design**

**Table S2. MPRA expression data**

**Table S3. MPRA fold change data**

**Table S4. Significance analysis of HPO term overlap**

**Table S5. RNA-seq fold change data**

**Table S6. PCR primers and conditions**

